# Continued and Serious Lockdown Could Minimize Many Newly Transmitted Cases of COVID-19 in the U.S.: Wavelets, Deterministic Models, and Data

**DOI:** 10.1101/2020.04.30.20080978

**Authors:** Arni S.R. Srinivasa Rao, Steven G. Krantz

## Abstract

All the newly reported COVID-19 cases of April in the U.S. have not acquired the virus in the same month. We estimate that there was an average of 29,000/day COVID-19 cases in the U.S. transmitted from infected to susceptible during April 1–24, 2020 after adjusting for under-reported and under-diagnosed. We have provided model-based predictions of COVID-19 for the low and high range of transmission rates and with varying degrees of preventive measures including the lockdowns. We predict that even if 10% of the susceptible and 20 % of the infected who were not identified as of April 23, 2020, do not adhere to proper care or do not obey lockdown, then by the end of May and by end of June 50,000 and 55,000 new cases will emerge, respectively. These values for the months of May and June with worse adherence rates of 50% by susceptible and infected (but not identified) will be 251,000 and 511,000, respectively. Continued and serious lockdown measures could bring this average daily new cases to a further low at 4,300/day to 8,000/day in May.

## Introduction

As the cumulative number of novel coronavirus cases in the U.S. is still very high at more than 895,766 as of April 24, 2020, and more than 709,665 of these cases were reported during April 1–24, 2020 [1], we have asked a couple of questions: namely, did the lockdown implemented by various states in the U.S. by the end of March 2020 have any impact so far, especially in preventing the new COVID-19 cases in the recent weeks in the U.S.? Will there be any degree of damage in further controlling the spread of COVID-19 in the U.S. by relaxing the lockdown measures to keep-up the economy? We found that relaxing ongoing efforts on controlling the pandemic in the U.S. could have striking impact on the number of new cases for the months of May and June 2020. A 50% relaxed approach in the lockdown or poor preventive measures in the U.S. could lead to additional new COVID cases up to 200,000 to 370,000. We provide several other scenarios too which are in Table 1 to Table 6.

**Table 1.** Predicted COVID-19 cases in the U.S. under the varying degrees of adhering to lockdown and other preventive measures from May 1 to June 30, 2020 (Medium range)

| End of the Month in 2020 | Predicted New COVID-19 Cases by Age groups (Medium Range)<br>(with percentage of people among Infected but not yet detected who are not adhering to lockdown and other preventive measures is = 20) |  |  |  | Percentage of susceptible people not adhering to Lockdown and other preventive precautions |
| --- | --- | --- | --- | --- | --- |
|  | <18 | 18-64 | 65+ | Total |  |
| **May** | 259 | 37,876 | 12,275 | 50,410 | 10 |
| **June** | 312 | 41,275 | 13,701 | 55,288 |  |
| **May** | 326 | 42,123 | 14,054 | 56,503 | 20 |
| **June** | 455 | 50,215 | 17,407 | 68,077 |  |
| **May** | 402 | 46,929 | 16,048 | 63,379 | 30 |
| **June** | 605 | 61,294 | 21,971 | 83,870 |  |
| **May** | 489 | 52,340 | 18,285 | 71,114 | 40 |
| **June** | 860 | 75,018 | 27,600 | 103,478 |  |
| **May** | 588 | 58,432 | 20,796 | 79,816 | 50 |
| **June** | 1,142 | 92,011 | 34,542 | 127,695 |  |
| **May** | 700 | 65,289 | 23,614 | 89,603 | 60 |
| **June** | 1,493 | 113,043 | 43,111 | 157,647 |  |
| **May** | 827 | 73007 | 26778 | 100,612 | 70 |
| **June** | 1931 | 139067 | 53690 | 194,688 |  |

**Table 2.** Predicted COVID-19 cases in the U.S. under the varying degrees of adhering to lockdown and other preventive measures from May 1 to June 30, 2020 (High range)

| End of the Month in 2020 | Predicted New COVID-19 Cases by Age groups (High Range)<br>(with percentage of people among Infected but not yet detected who are not adhering lockdown and other preventive measures is = 20) |  |  |  | Percentage of susceptible people not adhering to Lockdown and other preventive precautions |
| --- | --- | --- | --- | --- | --- |
|  | <18 | 18-64 | 65+ | Total |  |
| **May** | 315 | 38,012 | 13,918 | 52,245 | 10 |
| **June** | 430 | 41,961 | 16,923 | 59,314 |  |
| **May** | 460 | 42,991 | 17,684 | 61,135 | 20 |
| **June** | 775 | 53,741 | 25,234 | 79,750 |  |
| **May** | 642 | 49,186 | 22,104 | 71,932 | 30 |
| **June** | 1,277 | 70,781 | 36,482 | 108,540 |  |
| **May** | 866 | 56,818 | 27,315 | 84,999 | 40 |
| **June** | 1,992 | 95,007 | 51,854 | 148,853 |  |
| **May** | 1,140 | 66,151 | 33,484 | 100,775 | 50 |
| **June** | 3,002 | 129,108 | 72,982 | 205,092 |  |
| **May** | 1,475 | 77,507 | 40,807 | 119,789 | 60 |
| **June** | 4,418 | 176,827 | 102,126 | 283,371 |  |
| **May** | 1,882 | 91,270 | 49,521 | 142,673 | 70 |
| **June** | 6,396 | 243,366 | 142,408 | 392,170 |  |

**Table 3.** Predicted COVID-19 cases in the U.S. under the varying degrees of adhering to lockdown and other preventive measures from May 1 to June 30, 2020 (Low range)

| End of the Month in 2020 | Predicted New COVID-19 Cases by Age groups (Low Range)<br>(with percentage of people among Infected but not yet detected who are not adhering to lockdown and other preventive measures is = 50) |  |  |  | Percentage of susceptible people not adhering to Lockdown and other preventive precautions |
| --- | --- | --- | --- | --- | --- |
|  | <18 | 18-64 | 65+ | Total |  |
| **May** | 647 | 94,637 | 30,687 | 125,971 | 10 |
| **June** | 781 | 103,182 | 34,248 | 138,211 |  |
| **May** | 815 | 105,303 | 35,131 | 141,249 | 20 |
| **June** | 1,138 | 125,521 | 43,506 | 170,165 |  |
| **May** | 1,006 | 117,315 | 40,115 | 158,436 | 30 |
| **June** | 1,587 | 153,203 | 54,908 | 209,698 |  |
| **May** | 1,223 | 130,840 | 45,705 | 177,768 | 40 |
| **June** | 2,150 | 187,486 | 68,962 | 258,598 |  |
| **May** | 1,470 | 146,065 | 51,979 | 199,514 | 50 |
| **June** | 2,854 | 229,525 | 86,295 | 318,674 |  |
| **May** | 1,750 | 163,201 | 59,020 | 223,971 | 60 |
| **June** | 3,732 | 282,440 | 107,679 | 393,851 |  |
| **May** | 2,067 | 182,486 | 66,925 | 251,478 | 70 |
| **June** | 4,824 | 347,401 | 134,072 | 486,297 |  |

**Table 4.** Predicted COVID-19 cases in the U.S. under the varying degrees of adhering to lockdown and other preventive measures from May 1 to June 30, 2020 (High range)

| End of the Month in 2020 | Predicted New COVID-19 Cases by Age groups (High Range)<br>(with percentage of people among Infected but not yet detected who are not adhering to lockdown and other preventive measures is = 50) |  |  |  | Percentage of susceptible people not adhering to Lockdown and other preventive precautions |
| --- | --- | --- | --- | --- | --- |
|  | <18 | 18-64 | 65+ | Total |  |
| **May** | 787 | 95,029 | 34,791 | 130,607 | 10 |
| **June** | 1,075 | 104,893 | 42,290 | 148,258 |  |
| **May** | 1,150 | 107,472 | 44,199 | 152,821 | 20 |
| **June** | 1,937 | 134,318 | 63,033 | 199,288 |  |
| **May** | 1,604 | 122,953 | 55,238 | 179,795 | 30 |
| **June** | 3,189 | 176,861 | 91,092 | 271,142 |  |
| **May** | 2,164 | 142,020 | 68,253 | 212,437 | 40 |
| **June** | 4,975 | 237,308 | 129,403 | 371,686 |  |
| **May** | 2,850 | 165,336 | 83,657 | 251,843 | 50 |
| **June** | 7,493 | 322,333 | 182,010 | 511,836 |  |
| **May** | 3,683 | 193,699 | 101,938 | 299,320 | 60 |
| **June** | 11,020 | 441,204 | 254,480 | 706,704 |  |
| **May** | 4,702 | 228,070 | 123,685 | 356,457 | 70 |
| **June** | 159,40 | 606,754 | 354,478 | 961,232 |  |

**Table 5.** Predicted COVID-19 cases in the U.S. under the varying degrees of adhering to lockdown and other preventive measures from May 1 to June 30, 2020 (Medium range)

| End of the Month in 2020 | Predicted New COVID-19 Cases by Age groups (Medium Range) |  |  |  | Percentage of susceptible people not adhering to Lockdown and other preventive precautions |
| --- | --- | --- | --- | --- | --- |
|  | <18 | 18-64 | 65+ | Total |  |
| **May** | 1293 | 189270 | 61370 | 251933 | 10 |
| **June** | 1562 | 206345 | 68482 | 276389 |  |
| **May** | 1629 | 210594 | 70254 | 282477 | 20 |
| **June** | 2274 | 250987 | 86976 | 340237 |  |
| **May** | 2012 | 234608 | 80215 | 316835 | 30 |
| **June** | 3172 | 306295 | 109746 | 419213 |  |
| **May** | 2447 | 261664 | 91388 | 355499 | 40 |
| **June** | 4297 | 374772 | 137801 | 516870 |  |
| **May** | 2940 | 292078 | 103924 | 398942 | 50 |
| **June** | 5703 | 459510 | 172386 | 637599 |  |
| **May** | 3500 | 326328 | 117993 | 447821 | 60 |
| **June** | 7455 | 564324 | 215034 | 786813 |  |
| **May** | 4133 | 364869 | 133786 | 502788 | 70 |
| **June** | 9636 | 693920 | 267638 | 971194 |  |

**Table 6.** Predicted COVID-19 cases in the U.S. under the varying degrees of adhering to lockdown and other preventive measures from May 1 to June 30, 2020 (High range)

| End of the Month in 2020 | Predicted New COVID-19 Cases by Age groups (High Range) |  |  |  | Percentage of susceptible people not adhering to Lockdown and other preventive precautions |
| --- | --- | --- | --- | --- | --- |
|  | <18 | 18-64 | 65+ | Total |  |
| **May** | 1,573 | 190,051 | 69,566 | 261,190 | 10 |
| **June** | 2,148 | 209,755 | 84,521 | 296,424 |  |
| **May** | 2,300 | 214,924 | 88,360 | 305,584 | 20 |
| **June** | 3,870 | 268,519 | 125,895 | 398,284 |  |
| **May** | 3,206 | 245,862 | 110,407 | 359,475 | 30 |
| **June** | 6,370 | 353,414 | 181,803 | 541,587 |  |
| **May** | 4,325 | 283,959 | 136,393 | 424,677 | 40 |
| **June** | 9,929 | 473,922 | 258,039 | 741,890 |  |
| **May** | 5,696 | 330,535 | 167,137 | 503,368 | 50 |
| **June** | 14,942 | 643,223 | 362,546 | 1,020,711 |  |
| **May** | 7,366 | 387,176 | 203,612 | 598,154 | 60 |
| **June** | 21,954 | 879,549 | 506,202 | 1,407,715 |  |
| **May** | 9,393 | 455,793 | 246,982 | 712,168 | 70 |
| **June** | 31,716 | 1,208,000 | 703,876 | 1,943,592 |  |

It is not always easy to conduct experiments to get the parameters of transmissions during the lockdowns [2]. One can indirectly obtain the number of people who were infected in a geographical area within a specified time and obtain population-level incidence rates based on the newly reported cases. But we know that often not all cases are reported or diagnosed or both [3]. That means a true incidence rate in the population is not easy to obtain unless we adjust the reported cases with the number not reported and retrospectively adjust the past reported data [3].

## Methods, Models and Data

We want to understand through simple transmission dynamics principles what fraction of these cases were possibly acquired during April. We want to understand these numbers by adjusting for under-reporting including the under-diagnosis during the period. Further, we develop age-structured population models of COVId-19 spread and, combining the outcomes with wavelet analysis, we have provided impact of not adhering to lockdown and other preventive measures combined in the U.S. for the period May-June, 2020. Our methods, models and the data are described in the next paragraphs.

**Figure 1.**
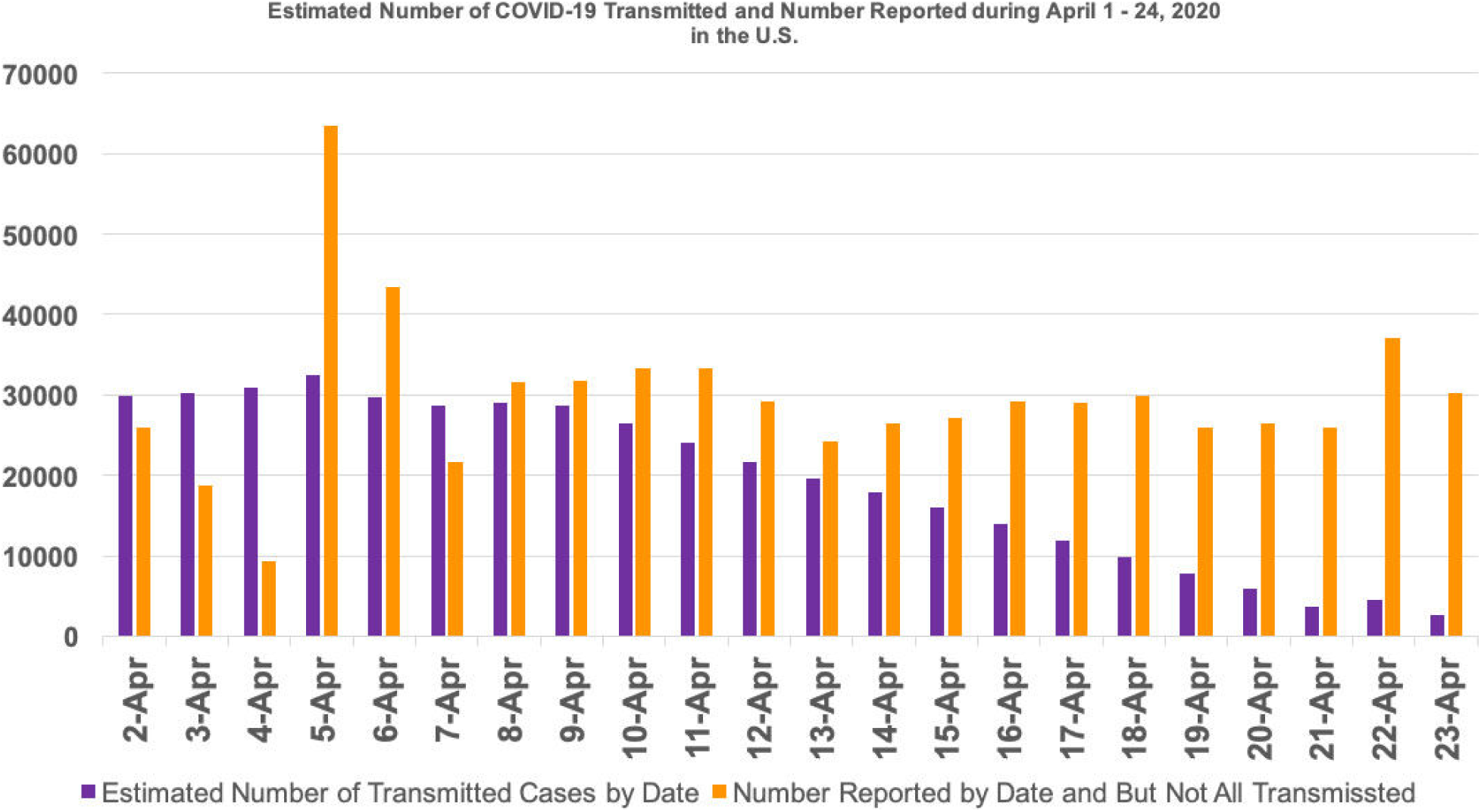
Estimated Transmissions occurred during each day and the number of reported per day during April 1–23, 2020. The estimated cases are skewed because some of the transmission occurred before April 19 might be reported at a later date.

Let *f_n_* be the cumulative number of COVID-19 cases reported at the end of the *n^th^* day and *f*_0_ = 0 be the number of COVID-19 cases at the beginning. Let *f_1_* be the number of cases at the end of *1^st^* day. Let *I_i_ = f_i+_*_1_ *− f_i_* for *i* = 0,1,2,…,*n* − 1 be the new cases on the each day. Let *c_i_* ∊ (0,1) be the fraction of *I_i_* those are infected on the *i^th^* day such that

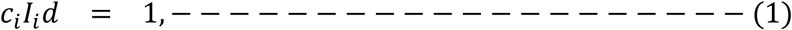

where *d* is the average incubation period. That is,

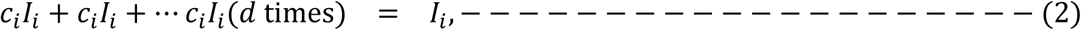

is satisfied. Through the equation (A.1), we have distributed newly reported cases on the *i^th^* day into *i^th^* day and *d* − 1 number of days prior to *i^th^* day. We assumed that *c_i_* is constant in (1), so that, *I_i_* is uniformly distributed during [*i*^th^ day, *d* days prior]. Instead of uniform distribution of *I_i_* as considered in (1), one can consider a skewed or non-uniform or some random distribution if there is strong evidence of the same. Under such non-uniform situations as well we will have 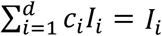, and 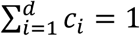. Once each *I_i_* value is partitioned into *d* − 1 days prior to the *i^th^* day, then the number of transmuted cases in each day is computed using the below set of equations:

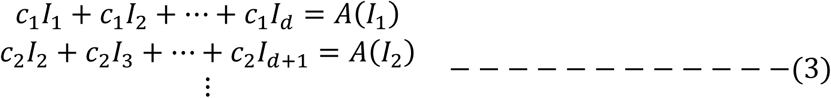

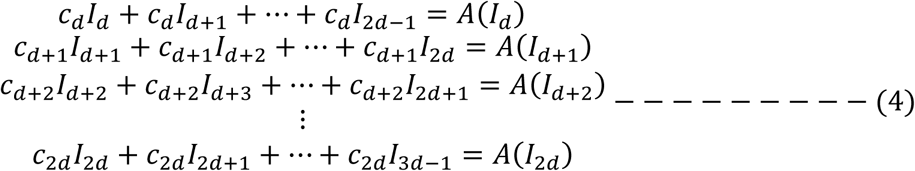

These continue until the last reported cases are available for a given population. For the computations in this article we considered uniform *c_i_* values over *d* days by assuming *c_i_ = c* within the period of study. Then we assumed that the daily new cases April 1 to April 13 were uniformly distributed into 1/14th, 2/14th, 3/14th and so on until 13/14th fractions of the reported and we assumed all the cases from April 14 were the result of those transmitted during the month of April. A similar backward adjustment of reported cases was theoretically demonstrated in our earlier article with advanced network structures [5].

Let *Y* be the total number of newly infected cases who were not traced into the system. Let *Y = Y*_1_ *+ Y*_2_*+Y*_3_, where *Y_k_* is the size of infected in the ag-group *k* for *k* =0–17, 18–64, 65+, respectively. Let *X* be the total number of susceptible individuals and 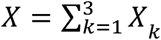, where *X_k_* is the size of susceptible in the ag-group *k*. Let *L(X_k_)* = Ø*_k_X_k_* be the fraction Ø*_k_* of susceptible *X_k_* who do not practice lockdown or other preventive measures in the age-group *k*, such that

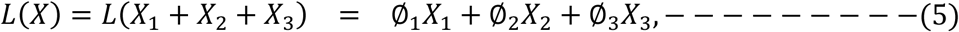

where 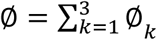 is the total fraction of susceptibles in the population who do not adhere to any preventive measures including the lockdown. Similarly, we will assume that

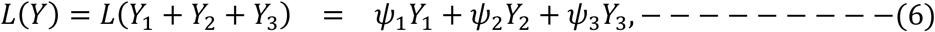

where 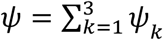 is the total fraction of infected (who are not traced) in the population who do not adhere to any preventive measures including the lockdown. The differential equations describing the transmission dynamics from infected to susceptible are given below:

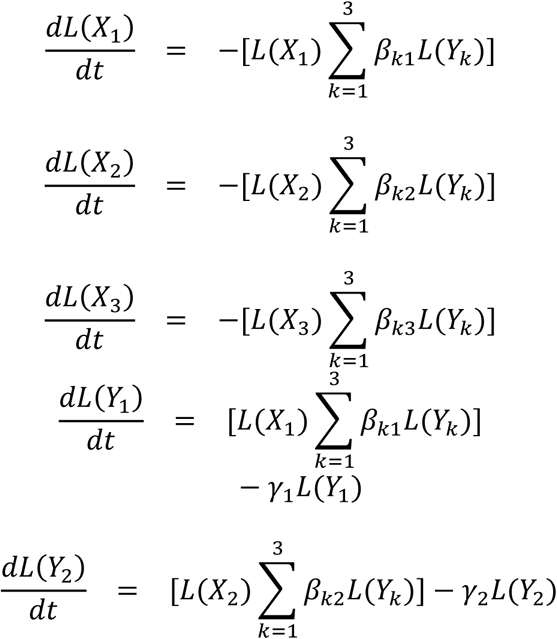

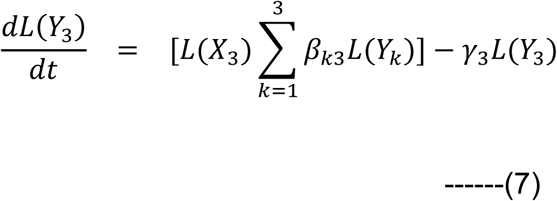

Here *β_kl_* is the average transmission rate from an infected in the age group *k* to a susceptible in the age group *l* and *γ_k_* is the recovery rate for the age-group *k*. Similar age structure population models for the COVID-19 were developed by us in predicting the U.S. population aged 65+ with underlying medical conditions [7]. Once we obtain predicted values of new COVID-19 cases we will use Meyer’s wavelets to demonstrate the difference of magnitudes between observed and predicted cases and wavelets for the difference between various degrees of not adhering to lockdown or other preventive measures. Meyer’s wavelets are a natural improvement over the Fourier series and transforms. A Fourier series *f*(*x*) is written in terms of coefficients *a*_0_, *a_n_* and *b_n_* for *n* ∈ Ν, the set of natural numbers, and with trigonometric functions as below

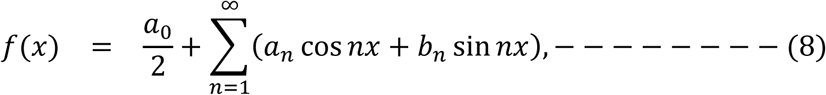

where cos *nx + i* sin *nx = e^inx^*. Further, if *f(x)* is Lebesgue integrable on an interval [0, *a*] with a period *a*, then we write

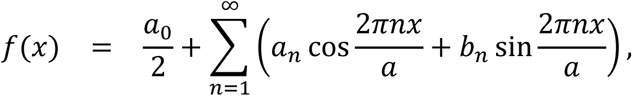

where

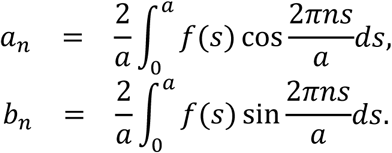

Once we have Fourier series, one can obtain Fourier transformations, *f_T_*, through (9), written as

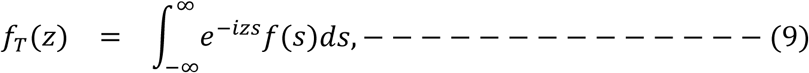

where *z* is a complex numbers and *f(s)* is a Fourier series function (8).

The Meyer wavelets *ψ(ω)* also use trigonometric functions which are infinitely differentiable on a certain domain. Several useful resources on wavelets are available in [8–14]. We have used such wavelets in COVID-19 modeling [3,5,7,15]. These wavelets when plotted helps us to distinguish the degree of difference in the magnitude between various levels of relaxation of lockdown and other preventive measures during COVID-19 lockdowns. The Meyer wavelets *U(ω)* together with accompanying function *ν* are given below.

What is interesting about this new wavelet technique is the following. The traditional trigonometric functions sine and cosine do not localize well. Once we know a trigonometric function on a small interval then it is uniquely determined on the entire real line. But wavelets localize nicely in both the space and phase variables. This makes them particularly useful for image compression, signal processing, and data analysis. Wavelets are the key to our new approach to understanding the corona pandemic.

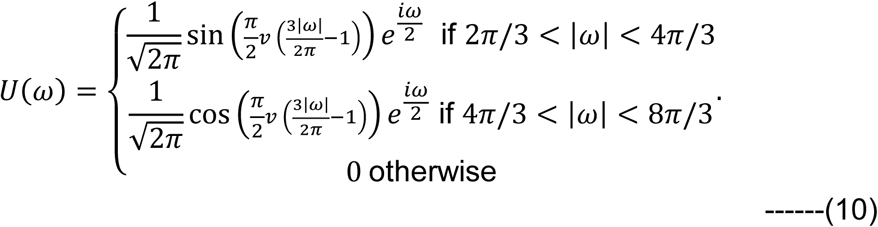

Here *ν*(*x*) = 0 for *x* < 0, and

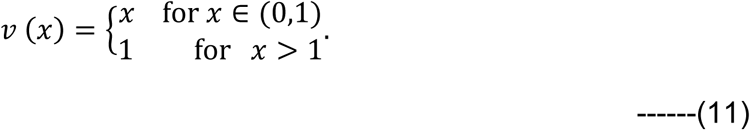

General population age-structure models are well-known in epidemiology, population biology, ecology, see for example [16–25]. However, the novelty of our work lies in linking the wavelets in conceptualizing the differences between the magnitudes of *L(Y*_1_*), [L(Y*_2_*), L(Y*_3_) with varying Ø and ψ values. Such exposure of Meyer’s wavelets or other wavelets were done earlier. For a more general application potential and beauty of applicability of wavelets one can refer to [31].

### Data

As of April 23, 2020 the age distribution of the COVID-19 cases in the U.S. among those whose age can be ascertained are as follows: 2,791 (aged <18), 475,659 (aged 18–64), 149,243 (aged 65+). The number of infected during April 1–22, 2020 who were not reported are estimated at 224,785 by our previous calculation of under-reporting in the U.S. [6]. These new cases of April we have divided into the age groups as per the CDC data above. This would provide us *Y*_1_ = 999, *Y*_2_ = 170,339, and *Y*_3_ = 53,446. Demographic data from census.gov [26] and adjusting it for annual growth that was observed over the past decadal growth gives us susceptible populations *X*_1_*, X*_2_*, X*_3_. We considered the lack of adherence from 10% to 70% for both susceptible and infected. We could provide combinations of these and provide more predictions, but we prepared for these finite number of combinations. One can try all other permutations and combinations. Distributing Ø and ψ within each age-groups, we have considered the following ratio [10%: 60%: 30%]. The β values for low range of predictions considered are

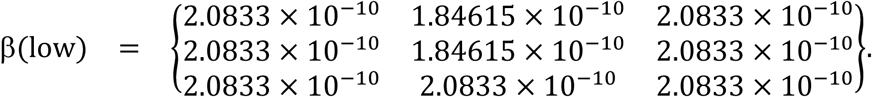

These low range of predictions are calibrated just as in our previous medium range of values in the study [7]. Originally, these transmission rates were calibrated earlier for the U.S. data on reported COVID-19 cases during March 1–14 and March 15-April 6, 2020 which were calibrated from our paper [3]. The high range transmission parameters considered are,

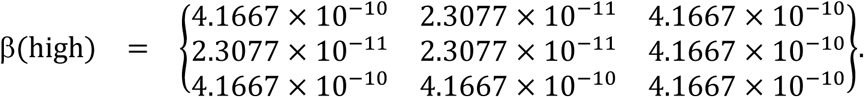

## Results

Out of the 709,665 COVID-19 cases newly reported during April 1–24, 2020, we have estimated that 560,524 cases were the result of transmissions within this period. To do this we have assumed a few things, the first being the average incubation period as per CDC as 2–14 days [4]. Adjusting the reported cases with the not-reported cases computed as of April 6, 2020, in the ratio of 1:1.5 as noted in [6] the true acquired estimated numbers during April 1–19 stands at 674,354. According to the CDC data released on April 23, 23% of the cases are in the age group 65+ [1]. This implies that 23% of 551,630, that is, 155,101 number of new cases who are aged 65+ occurred during the period. This number is consistent with another modeling study on 65+ [7] which predicts the number of new cases during April – June 2020.

**Figure 2.**
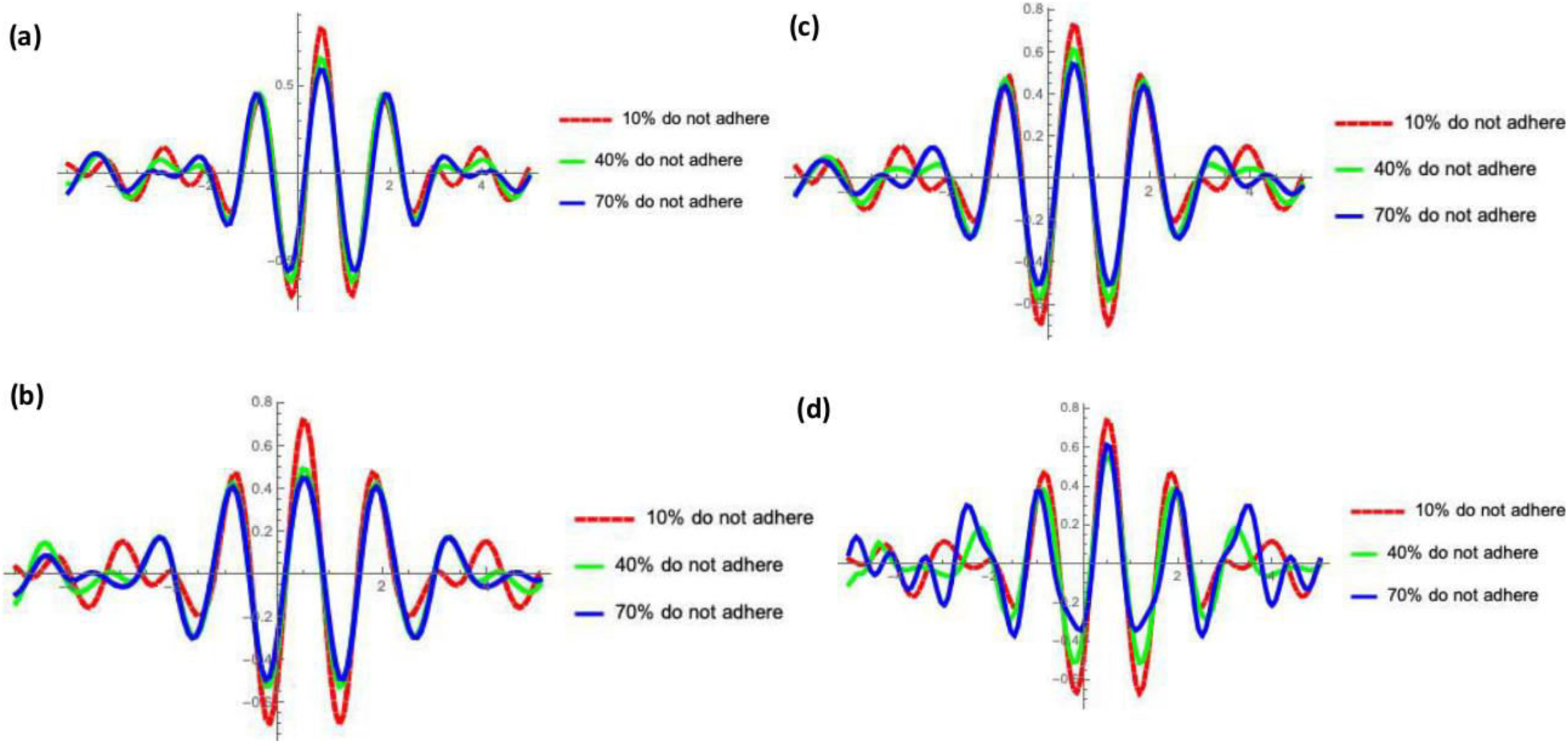
Meyer wavelets for the difference between magnitudes of predicted new COVID-19 cases in the U.S. for the months of May and June, 2020. All four sets are drawn based on ψ = 20% with ∅ = 10%, 40%, 70%. We have provided details of terminology and methods adopted in the Appendix and actual predicted values are in Tables 1 through Table 6. (a) Difference of magnitude in the month of May for Low range predictions, (b) Difference of magnitude in the month of June for Low range predictions, (c) Difference of magnitude in the month of May for High range predictions, (d) Difference of magnitude in the month of June for High range predictions.

In-addition to backward computation of newly transmitted cases in the month o April 2020 explained above and also in the Appendix, we have also developed dynamics models to understand the spread of COVID-19 for the immediate months in the U.S. We further used wavelets to demonstrate the difference in the spread that could occur due to varying degrees of seriousness of the lockdown implementation by susceptible and infected (who are not aware of their status). We assumed all the detected cases are been either self-quarantined or adhering to the preventive measures.

Serious preventive measures and continued lockdown could bring down the new cases in May at In-addition to the main results of our study mentioned in the introduction. We also note that relaxed approach towards lockdown and other preventive measures might be a burden on the hospital system.

**Figure 3.**
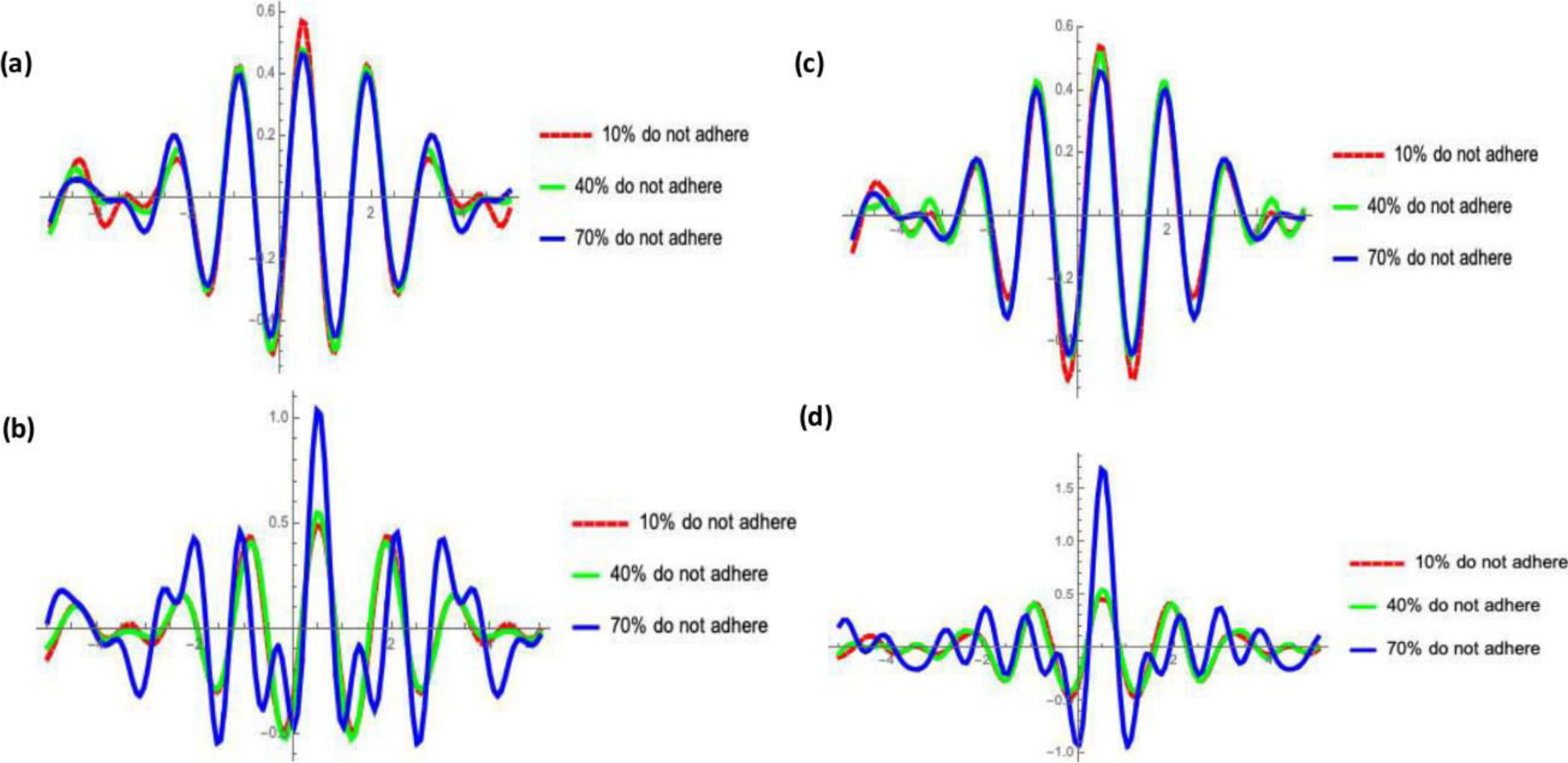
Meyer wavelets for the difference between magnitudes of predicted new COVID-19 cases in the U.S. for the months of May and June, 2020. All the four sets are drawn based on ψ = 50% with Ø = 10%, 40%, 70%. (a) Difference of magnitude in the month of May for Low range predictions, (b) Difference of magnitude in the month of June for Low range predictions, (c) Difference of magnitude in the month of May for High range predictions, (d) Difference of magnitude in the month of June for High range predictions.

## Concluding Remarks

In summary, we see that on average about 29,000/day new cases occurred that were transmitted during April 1–23, 2020. This is very high. This possibly indicates that the lockdowns and prevention measures have not been adhered to at the fullest. A complete lockdown could bring new infections to a much lower level in the next few weeks. Average daily cases with strict measures can be brought down to 4,300/day to 8,000/day in May. There is reason to believe that under-reporting is skewing our understanding of coronavirus infection and spread. It can be shown that lockdown measures are among the most effective for controlling virus development and spread.

As an ending remark, we want to emphasize that one could build more complex models for the transmission dynamics of COVID-19 in the US or for any population if there is strong evidence of transmission level parameters at the sub-population level. Making models more complex unnecessarily without any supportive evidence would not yield any better results and sometimes such exercises could bring misleading results [33]. There are two levels of complexities one can introduce, one at the level of the parameters and another at the subpopulation selection level but that needs very careful assessment of the situation, and modeling should not stand as a mere mathematical exercise.

Premature lifting of the lockdown could lead to disaster.

## Data Availability

Yes

## Authors contributions

Both the authors contributed in writing. ASRS Rao designed the study, developed the methods, models, collected data, performed analysis, computing, creating Figures and Tables, wrote the first draft. SG Krantz participated in the design, interpretation of results, and contributed in editing the draft, approving the methods. Both the authors approved the final manuscript.

